# Distinct medical and substance use histories associate with cognitive decline in Alzheimer’s Disease

**DOI:** 10.1101/2024.11.26.24317918

**Authors:** Clayton Mansel, Diego R. Mazzotti, Ryan Townley, Mihaela E Sardiu, Russell H. Swerdlow, Robyn A Honea, Olivia J Veatch

## Abstract

**INTRODUCTION:** Phenotype clustering reduces patient heterogeneity and could be useful when designing precision clinical trials. We hypothesized that the onset of early cognitive decline in patients would exhibit variance predicated on the clinical history documented prior to an Alzheimer’s Disease (AD) diagnosis

**METHODS:** Self-reported medical and substance use history (i.e., problem history) was used to cluster participants from the National Alzheimer’s Coordinating Centers (NACC) into distinct subtypes. Linear mixed effects modeling was used to determine the effect of problem history subtype on cognitive decline over two years.

**RESULTS:** 2754 individuals were partitioned into three subtypes: minimal (n = 1380), substance use (n = 1038), and cardiovascular (n = 336) subtypes. The cardiovascular problem history subtype had significantly worse cognitive decline over a two-year follow-up period (p = 0.013).

**DISCUSSION:** Our study highlights the need to account for problem history to reduce heterogeneity of outcomes in AD clinical trials.

## 1. Background

Alzheimer’s disease (AD) is the most common form of dementia, with an estimated prevalence in the U.S. of 6 million as of 2020 [1]. AD presentation is clinically and biologically heterogeneous with many factors affecting the progression of the disease including socioeconomic status [2], nutrition [3], apolipoprotein E (*APO3*) genotype [4], sex [4], and co-morbidities including diabetes, depression, and hypertension [5–7]. Decades of clinical trials have resulted in only three Food and Drug Administration approved therapies for AD in the last 20 years which showed only a modest slowing of cognitive decline [8,9]. One reason for these underwhelming results may be that AD trials generally enroll participants as one homogenous group, despite evidence that AD progression and response to therapy is highly heterogeneous [10]. When not accounted for, this heterogeneity could inflate the type II error rate in clinical trials and minimize the average treatment effect of interventions when relevant subgroups are not accounted for.

The advent of large biomedical databases and machine learning algorithms, such as clustering, enables precision medicine approaches focused on identifying subgroups of individuals according to complex patterns in a hypothesis-independent manner and reducing heterogeneity in patient populations [11–13]. These data-driven clusters can then be used to augment future clinical trial enrollment and predict treatment response [14,15]. For example, Seymour and colleagues [16] used 29 variables to retrospectively cluster participants in the ProCESS clinical trial which aimed to improve outcomes in patients with sepsis [17]. Even though the original trial showed nearly zero percent chance of benefit, when informed by phenotype clusters derived in this study, the chance of benefit rose to 35% [16].

Most previous clustering studies in AD utilize either cognitive tests [18–21] or biological data such as neuroimaging or fluid biomarkers in patients after a diagnosis has been made [22–24]. However, when using these data, it can be difficult to distinguish between true subgroups or different stages of the disease after onset [25]. Moreover, clinical trials generally try to target individuals early in the disease progression [8,9] when cognitive decline and biomarkers are less pronounced. One approach to address this dilemma is to use medical and substance use history (i.e., problem history) prior to disease onset which is commonly collected at every routine primary care visit. We hypothesized that problem history subtypes could be especially relevant to AD heterogeneity given that many previous studies have shown that co-morbidities and substance use, such as smoking and alcohol intake, affect AD progression [6,26,27]. A few previous studies have performed clustering in AD using problem history data; however, they utilize data from the electronic health record (EHR) [28–31]. The accuracy of AD diagnoses using International Classification of Diseases (ICD) billing codes is mixed at best [32–35] with one study identifying a lack of cognitive testing and time as major barriers in a primary care setting [36]. To ensure accurate AD diagnosis, we utilized the Uniform Data Set (UDS) [37], with consistent testing and diagnosis procedures, from the National Alzheimer’s Coordinating Centers (NACC) which is comprised of specialist Alzheimer’s Disease Research Centers (ADRCs) across the U.S. and is one of the largest clinical databases of individuals with AD in the world. In a cohort of individuals with AD, we performed multivariate clustering of medical and substance use survey items (problem history items) collected prior to AD diagnosis. To infer whether problem history subtypes could be used to inform future AD clinical trials, we compared each cluster’s mean trajectory of cognitive decline over the next two years.

## 2. Methods

### 2.1 Data source

This cross-sectional study analyzed data from UDS annual visits between September 2005 and November 2020 across 40 ADRCs in the NACC database. Details regarding data collection are well documented.[38] For individual problem history, self-reported data (available 2005-present) were chosen instead of clinician-assessed medical conditions (available 2015-present) to minimize missingness and increase statistical power.

29,818 individuals in the NACC database had at least 1 follow-up visit. Individuals were included if he or she had: 1) normal cognitive status, impaired but not mild cognitive impairment (MCI), or MCI at the initial visit, 2) dementia at any follow-up visit, and 3) dementia was determined to have a “primary etiology” of AD. The primary etiology of dementia was determined using the individual’s most recent visit to maximize diagnosis accuracy. Individuals <50 years old were removed to exclude autosomal-dominant forms of AD. Additionally, 298 individuals with missing problem history data were excluded. Ultimately, data from 2,754 individuals were included in the final cluster analysis (Figure 1; Table 1).

**Table 1.**
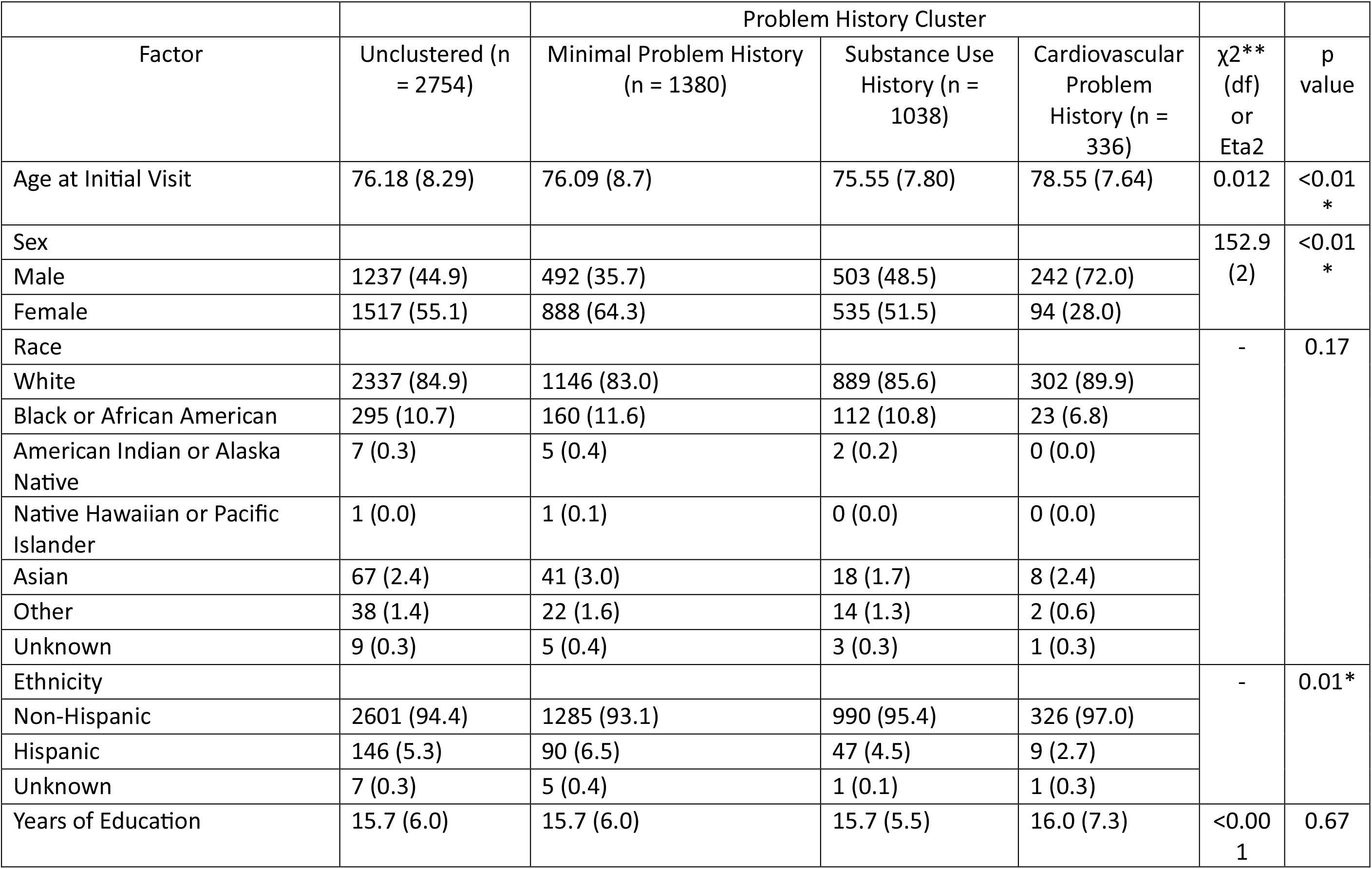
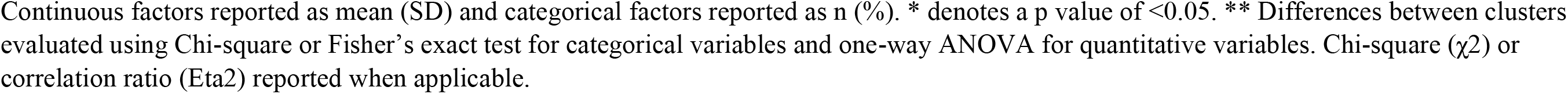
Demographics of NACC Cohort by Problem History Cluster.

| Factor | Unclustered (n = 2754) | Problem History Cluster | | | $\chi^2$**<br>(df)<br>or<br>Eta2 | p<br>value |
| --- | --- | --- | --- | --- | --- | --- |
|  |  | Minimal Problem History<br>(n = 1380) | Substance Use<br>History (n =<br>1038) | Cardiovascular<br>Problem<br>History (n =<br>336) |  |  |
| Age at Initial Visit | 76.18 (8.29) | 76.09 (8.7) | 75.55 (7.80) | 78.55 (7.64) | 0.012 | <0.01<br>* |
| Sex |  |  |  |  | 152.9<br>(2) | <0.01<br>* |
| Male | 1237 (44.9) | 492 (35.7) | 503 (48.5) | 242 (72.0) |  |  |
| Female | 1517 (55.1) | 888 (64.3) | 535 (51.5) | 94 (28.0) |  |  |
| Race |  |  |  |  | - | 0.17 |
| White | 2337 (84.9) | 1146 (83.0) | 889 (85.6) | 302 (89.9) |  |  |
| Black or African American | 295 (10.7) | 160 (11.6) | 112 (10.8) | 23 (6.8) |  |  |
| American Indian or Alaska<br>Native | 7 (0.3) | 5 (0.4) | 2 (0.2) | 0 (0.0) |  |  |
| Native Hawaiian or Pacific<br>Islander | 1 (0.0) | 1 (0.1) | 0 (0.0) | 0 (0.0) |  |  |
| Asian | 67 (2.4) | 41 (3.0) | 18 (1.7) | 8 (2.4) |  |  |
| Other | 38 (1.4) | 22 (1.6) | 14 (1.3) | 2 (0.6) |  |  |
| Unknown | 9 (0.3) | 5 (0.4) | 3 (0.3) | 1 (0.3) |  |  |
| Ethnicity |  |  |  |  | - | 0.01* |
| Non-Hispanic | 2601 (94.4) | 1285 (93.1) | 990 (95.4) | 326 (97.0) |  |  |
| Hispanic | 146 (5.3) | 90 (6.5) | 47 (4.5) | 9 (2.7) |  |  |
| Unknown | 7 (0.3) | 5 (0.4) | 1 (0.1) | 1 (0.3) |  |  |
| Years of Education | 15.7 (6.0) | 15.7 (6.0) | 15.7 (5.5) | 16.0 (7.3) | <0.00<br>1 | 0.67 |

Continuous factors reported as mean (SD) and categorical factors reported as n (%). \* denotes a p value of <0.05. \*\* Differences between clusters evaluated using Chi-square or Fisher's exact test for categorical variables and one-way ANOVA for quantitative variables. Chi-square ( $\chi^2$ ) or correlation ratio (Eta2) reported when applicable.

**Figure 1:**
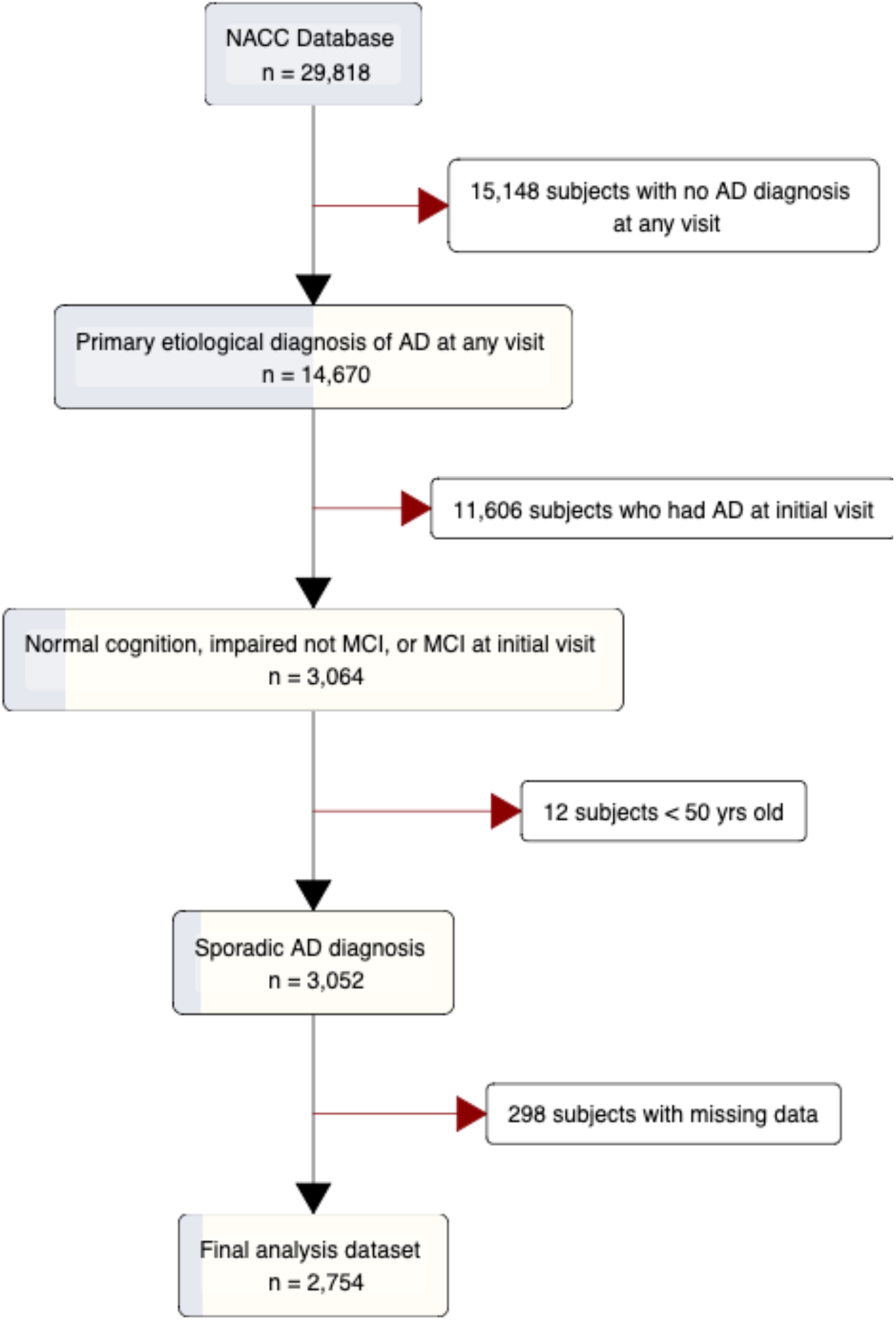
Flowchart of study cohort selection. NACC: National Alzheimer’s Coordinating Centers, AD: Alzheimer’s Disease, MCI: Mild Cognitive Impairment

### 2.2 Cluster Analysis

In total, 26 self-reported variables reflecting problem history were used as input for clustering (Table 2). Variables ranged from cardiovascular disease history to neurological conditions and substance use. For each variable, individuals could indicate “Absent”, “Recent/Active”, or “Remote/Inactive.” Both the “Recent/Active” and “Remote/Inactive” answers were collapsed into “Present” because 1) the difference was often not clinically relevant and 2) the individuals’ judgment of what is “remote” may introduce bias into the study.

**Table 2.** Problem History Items by Cluster.

| | Item | Unclustered<br>n = 2754 | Minimal<br>Problem<br>History<br>n = 1380 | Substance<br>Use<br>History<br>n = 1038 | Cardiovascul<br>ar Problem<br>History<br>n = 336 | $\chi^2$ (df) | p<br>value |
| --- | --- | --- | --- | --- | --- | --- | --- |
| Cardiovascular<br>Disease | Heart Attack/Cardiac Arrest | 169 (6.13) | 8 (0.58) | 17 (1.64) | 144 (42.86) | 897.03 (2) | <0.01<br>* |
|  | Atrial Fibrillation | 200 (7.26) | 59 (4.28) | 72 (6.94) | 69 (20.54) | 106.34 (2) | <0.01<br>* |
|  | Angioplasty/Endarterectomy/<br>Stent | 203 (7.37) | 5 (0.36) | 16 (1.54) | 182 (54.17) | 1228.58<br>(2) | <0.01<br>* |
|  | Cardiac Bypass Procedure | 137 (4.97) | 1 (0.07) | 2 (0.19) | 134 (39.88) | - | <0.01<br>* |
|  | Pacemaker | 95 (3.45) | 23 (1.67) | 16 (1.54) | 56 (16.67) | 200.76 (2) | <0.01<br>* |
|  | Congestive Heart Failure | 56 (2.03) | 5 (0.36) | 8 (0.77) | 43 (12.80) | 223.09 (2) | <0.01<br>* |
|  | Other Cardiovascular Disease | 289 (10.49) | 119<br>(8.62) | 120<br>(11.56) | 50 (14.88) | 13.28 (2) | <0.01<br>* |
|  | Diabetes | 334 (12.13) | 159<br>(11.52) | 112<br>(10.79) | 63 (18.75) | 16.05 (2) | <0.01<br>* |
|  | Hypertension | 1465 (53.20) | 690 (50) | 524<br>(50.48) | 251 (74.70) | 71.15 (2) | <0.01<br>* |
|  | Hypercholesterolemia | 1474 (53.52) | 651<br>(47.17) | 557<br>(53.66) | 266 (79.17) | 111.19 (2) | <0.01<br>* |
| Metabolic Conditions | Vitamin B12 Deficiency | 155 (5.62) | 79 (5.72) | 59 (5.68) | 17 (5.06) | 0.23 (2) | 0.89 |
|  | Thyroid Disease | 539 (19.57) | 280<br>(20.29) | 194<br>(18.69) | 65 (19.35) | 0.98 (2) | 0.61 |
| Neurological<br>Conditions | Stroke | 111 (4.03) | 57 (4.13) | 25 (2.41) | 29 (8.63) | 25.48 (2) | <0.01<br>* |
|  | Traumatic Brain Injury (TBI) | 273 (9.91) | 126<br>(9.13) | 116<br>(11.18) | 31 (9.23) | 2.98 (2) | 0.23 |
|  | Seizures | 51 (1.85) | 23 (1.67) | 20 (1.93) | 8 (2.38) | 0.81 (2) | 0.67 |
|  | Parkinson's Disease | 8 (0.29) | 8 (0.58) | 0 (0) | 0 (0) | - | 0.02* |
|  | Other Parkinsonian Disorder | 16 (0.58) | 8 (0.58) | 6 (0.58) | 2 (0.60) | - | 0.99 |
|  | Urinary Incontinence | 353 (12.82) | 176<br>(12.75) | 126<br>(12.14) | 51 (15.18) | 2.11 (2) | 0.35 |
|  | Fecal Incontinence | 72 (2.61) | 53 (3.84) | 13 (1.25) | 6 (1.79) | 16.62 (2) | <0.01<br>* |
| Psychiatric Disorders | Active Depression in the last<br>2 years | 821 (29.81) | 417<br>(30.22) | 315<br>(30.35) | 89 (26.49) | 2.02 (2) | 0.36 |
|  | Other Psychiatric Disorder | 128 (4.65) | 57 (4.13) | 62 (5.97) | 9 (2.68) | 7.89 (2) | 0.019<br>* |
| Tobacco Use | Smoked Cigarettes in the Last<br>30 Days | 85 (3.09) | 1 (0.07) | 72 (6.94) | 12 (3.57) | - | <0.01<br>* |
|  | Smoked More than 100<br>Cigarettes in Life | 1199 (43.54) | 20 (1.45) | 1027<br>(98.94) | 152 (45.24) | 2290.94<br>(2) | <0.01<br>* |
|  | Average Number of Packs<br>Smoked Per Day: |  |  |  |  | - | <0.01<br>* |
|  | 0 | 1552 (56.35) | 1360<br>(98.55) | 9 (0.87) | 183 (54.46) |  |  |
|  | 1 cigarette to < 1/2 pk | 417 (15.14) | 10 (0.72) | 367<br>(35.36) | 40 (11.90) |  |  |
|  | 1/2 pk to < 1 pk | 406 (14.74) | 9 (0.65) | 358<br>(34.49) | 39 (11.61) |  |  |
|  | 1 pk to 1 & 1/2 pks | 196 (7.12) | 0 (0) | 163<br>(15.70) | 33 (9.82) |  |  |
|  | 1 & 1/2 pks to 2 pks | 99 (3.59) | 0 (0) | 79 (7.61) | 20 (5.95) |  |  |
|  | > 2 pks | 84 (3.05) | 1 (0.07) | 62 (5.97) | 21 (6.25) |  |  |
| Substance Use | Alcohol Abuse - Clinically Significant | 98 (3.56) | 20 (1.45) | 67 (6.45) | 11 (3.27) | 43.34 (2) | <0.01 * |
|  | Other Substance Abuse | 13 (0.47) | 0 (0) | 13 (1.25) | 0 (0) | - | <0.01 * |

Variable categories (n (%)) shown for every variable used to construct the problem history clusters. Differences between the clusters were tested using a Chi-square test of independence. If the expected frequency was less than five, a Fisher's exact test was utilized. \* denotes a p value of <0.05.

Data preprocessing and clustering was conducted in R (version 4.3.1) and RStudio (version 2023.06.1+524). Initially, multiple correspondence analysis (MCA) [39] was performed to: 1) reduce the overall dimensionality of the data, and 2) transform categorical data into continuous component scores for clustering. To determine the number of components necessary to explain the majority of the variability observed in the dataset, the mean-squared error of prediction (MSEP) was plotted after performing k-fold cross-validation with 5% missing values added to the dataset across 100 simulations using the missMDA (v1.18) package in R. Five components substantially decreased the MSEP and were thus retained as input for clustering. The FactoMineR (v2.8) [40] and factoextra (v1.07) packages were used to perform MCA and visualize results.

Agglomerative hierarchical clustering was performed on individual component scores using the FactoMineR (v2.8) package. Individual similarity was determined using Euclidean distance and Ward’s method to build the tree [41]. Inertia gain estimates were calculated when dividing the dataset between 2 and 10 clusters. The final cluster solution was determined by the largest relative drop in inertia gain which resulted in three clusters [39]. See Supplementary Figure 1 for the dendrogram as well as the top five most closely associated categories for each problem history cluster. Overall, clusters characterized and named according to which problem history items were present in higher proportions than the other clusters (Table 2).

### 2.3 Supplemental Variable Definitions

Clusters were further characterized by other features not used as input in the clustering algorithm. Age, sex, race, and ethnicity were self-reported and provided by the NACC. *APOE* genotypes were supplied by the NACC when available [42–44]. The NACC received genotypes from the participating ADRCs, ADGC, and the National Centralized Repository for Alzheimer’s Disease (https://naccdata.org).

### 2.4 Statistics and Longitudinal Analysis

First, chi-square tests of independence (or Fisher’s exact test when the expected count of a category was n<5) were performed to determine differences in categorical non-transformed input variables and supplemental variables between clusters. One-way analyses of variance (ANOVA) were used to compare continuous supplemental measures among clusters. Unadjusted p-values less than 0.05 were considered significant.

The progression of cognitive decline for individuals in each cluster was characterized using a linear mixed effects model. The outcome variable was the Clinical Dementia Rating Sum of Boxes (CDRSUM) which ranges from 0 to 18, with higher numbers indicating worsening cognitive decline. CDRSUM was chosen because of its clinical relevance and use as an outcome in recent AD clinical trials [9]. Three annual study visits were included in the longitudinal analysis to: 1) minimize the effect of study dropout between subsequent visits and 2) focus on a follow-up period that is typical for a phase III AD clinical trial [8,9]. Age, sex, *APOE* genotype, baseline CDR, visit number, and cluster membership were included as fixed effects and subject ID was included as a random effect to account for repeated measures. Prior to conducting analyses, 316 (11.4%) individuals were dropped because *APOE* genotype was missing, leaving 2,438 individuals. Of these, 2,407 individuals (98.7%) completed at least three NACC study visits. To account for non-random censoring between clusters, the model was weighted by the inverse probability of censoring estimated using a binomial general linear model with the same fixed effects as the unweighted model. Sex by cluster and age by cluster interaction terms were tested in the weighted model and not included because they were not significant at p = 0.05.

A likelihood ratio test was used to evaluate the contribution of the problem history cluster on CDRSUM trajectory. To investigate differences between clusters at visit three, the pairwise contrasts between the marginal means of each cluster was compared using a Wald test. Unadjusted p values less than 0.05 were considered significant. Longitudinal analyses were conducted using R version 4.3.1 and the *lme4, ggeffects*, and *ipw* packages.

## 3. Results

Overall, there were 2,754 participants in the NACC database who did not have AD at their initial visit but were diagnosed at a later visit and met cohort criteria (see Methods and Figure 1). The overall dataset had a mean (SD) age of 76.2 (8.3) years, was 55.1% female, 84.9% white, 94.4% non-Hispanic, and had a mean (SD) of 15.7 (6.0) years of education (Table 1). We then performed hierarchical clustering on principal components from MCA, which resulted in three problem history subtypes summarized in Table 2. The three subtypes can be described as 1) minimal problem history (n = 1,380), 2) substance use history (n = 1,038), and 3) cardiovascular problem history (n = 336). Age at initial visit, sex, and ethnicity were significantly different across the three subtypes (Table 1). The cardiovascular problem history subtype had a numerically higher mean age of 78.6, lower proportion of females (28.0%), and higher proportion of non-Hispanic individuals (97.0%) (Table 1). The demographics of race and years of education were not significantly different across the subtypes. Every problem history item was significantly different between the subtypes except vitamin B12 deficiency, thyroid disease, traumatic brain injury, seizures, other Parkinsonian disorders, urinary incontinence, and depression (Table 2). The minimal problem history subtype was noted to have the most problem history variables reported as ‘Absent’ across nearly all categories, including fewer reports of smoking >100 cigarettes lifetime (1.5%), angioplasty/endarterectomy/stent placement (0.4%), and cardiac arrest (0.6%) (Table 2). The substance use history subtype had the most individuals indicating a smoking (6.9%) and alcohol abuse (6.5%) history compared to other subtypes (Table 2). The cardiovascular problem history subtype had the highest proportion of individuals indicating a significant cardiovascular disease history including a heart attack/cardiac arrest (42.9%), atrial fibrillation (20.5%), and hypertension (74.4%), among others (Table 2).

Next, we further characterized these problem history subtypes according to the age of AD diagnosis, family history, *APOE* genotype, and co-occurrence of other types of dementia. The age of AD diagnosis was significantly different across subtypes with the cardiovascular problem history cluster having the numerically highest mean age (80.9) (Table 3). It should be noted that the overall mean (SD) age at the NACC initial visit was 76.2 (8.3) and the overall mean (SD) age of AD diagnosis was 78.5 (8.8) which results in a mean (SD) difference of 2.4 (2.5) between evaluation and later diagnosis. The subtypes were also significantly different with respect to early-onset (age range 50 to 64) versus late-onset (>65 years) AD. The minimal problem history subtype had the numerically highest proportion of early-onset AD (6.6%) and the cardiovascular problem history subtype had numerically highest proportion of late-onset AD (97.0%) (Table 3). The proportion of a self-reported family history of AD was also significantly different on both the maternal and paternal side (Table 3). The cardiovascular problem history subtype had a markedly lower proportion of individuals reporting a maternal family history (28.9%) than the overall dataset (36.3%). The cardiovascular problem history subtype also had a numerically lower proportion of individuals reporting a paternal family history, although the comparison to the overall dataset was less (14.9% vs 17.1% for the cardiovascular problem history subtype compared to the overall dataset respectively). The difference in the frequency of *APOE* genotypes across subtypes was nearly significant (p = 0.08). Compared to the overall dataset, the largest differences with respect to *APOE* genotype were the ε3/ε3 genotype in the cardiovascular problem history subtype (36.0% vs 43.8%) and the ε4/ε4 genotype in the cardiovascular problem history subtype (9.8% vs 6.2%) in the overall dataset versus within-subtype respectively. No differences between subtypes were observed with respect to the co-occurrence of vascular, Lewy body, or frontotemporal dementia (Table 3).

**Table 3.**
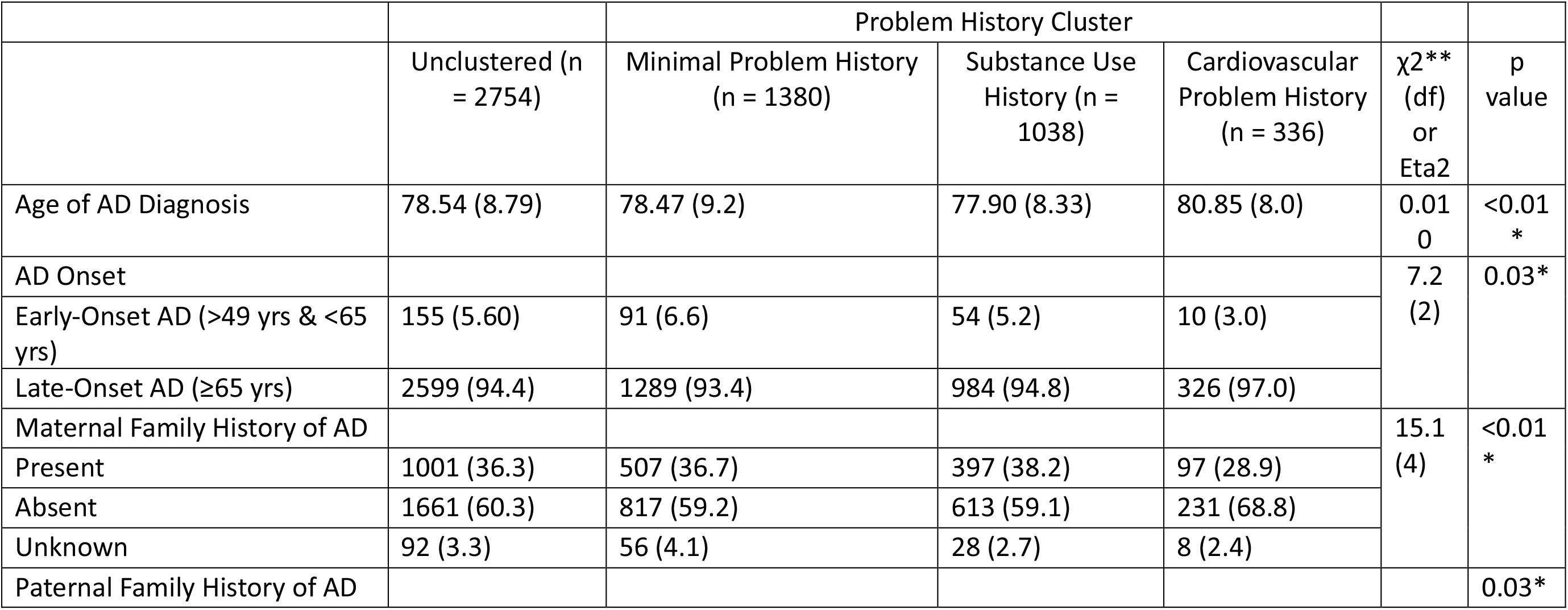
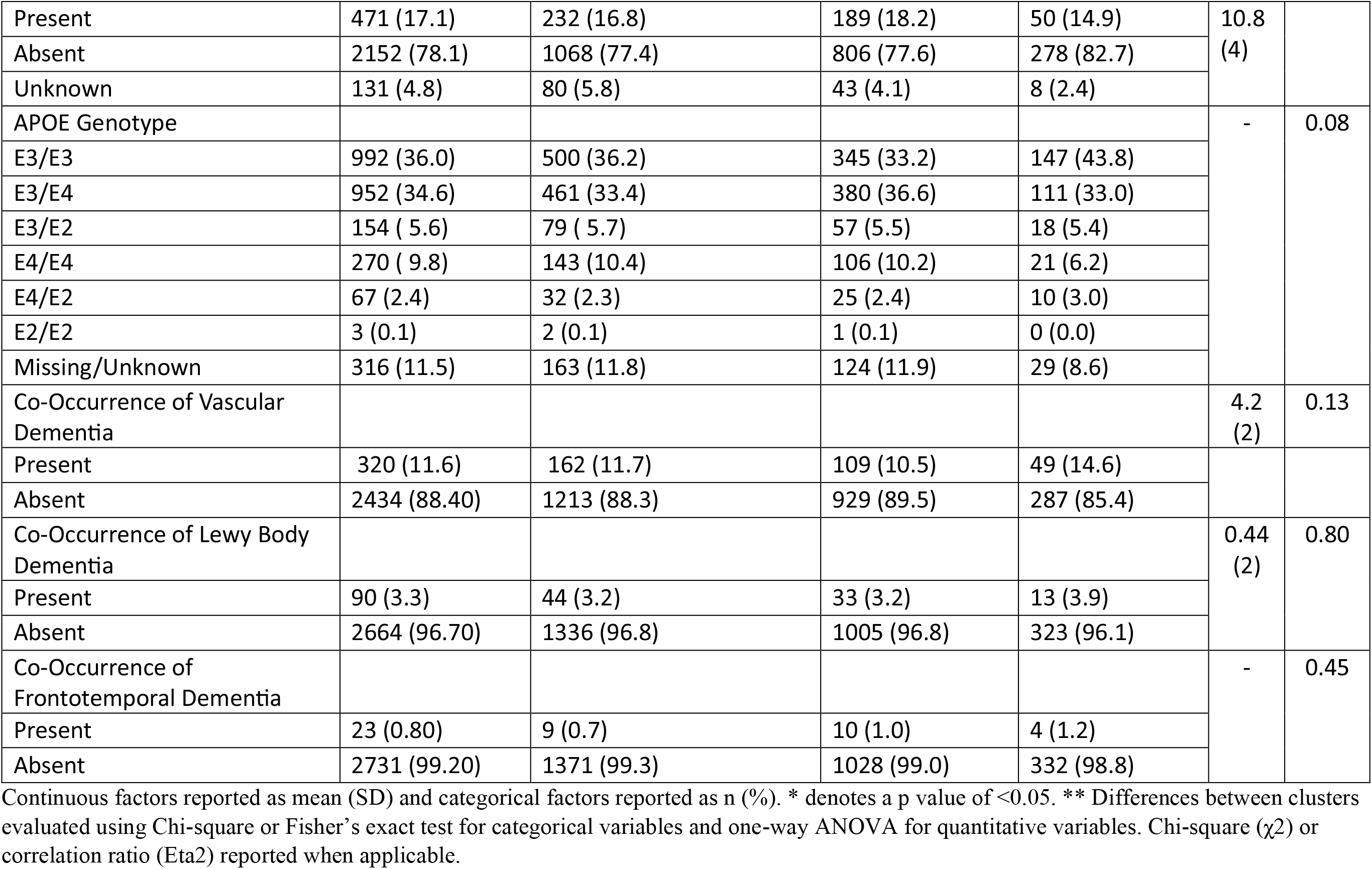
Alzheimer’s Disease Age of Onset, Family History, *APOE* Genotype, and Other Dementia Co-Occurrence for Each Problem History Cluster.

|  |  | Problem History Cluster |  |  |  |  |
| --- | --- | --- | --- | --- | --- | --- |
| | Unclustered (n = 2754) | Minimal Problem History (n = 1380) | Substance Use History (n = 1038) | Cardiovascular Problem History (n = 336) | $\chi^2$** (df) or Eta2 | p value |
| Age of AD Diagnosis | 78.54 (8.79) | 78.47 (9.2) | 77.90 (8.33) | 80.85 (8.0) | 0.010 | <0.01 * |
| AD Onset |  |  |  |  | 7.2 (2) | 0.03* |
| Early-Onset AD (>49 yrs & <65 yrs) | 155 (5.60) | 91 (6.6) | 54 (5.2) | 10 (3.0) |  |  |
| Late-Onset AD (≥65 yrs) | 2599 (94.4) | 1289 (93.4) | 984 (94.8) | 326 (97.0) | | |
| Maternal Family History of AD |  |  |  |  | 15.1 (4) | <0.01 * |
| Present | 1001 (36.3) | 507 (36.7) | 397 (38.2) | 97 (28.9) |  |  |
| Absent | 1661 (60.3) | 817 (59.2) | 613 (59.1) | 231 (68.8) |  |  |
| Unknown | 92 (3.3) | 56 (4.1) | 28 (2.7) | 8 (2.4) |  |  |
| Paternal Family History of AD |  |  |  |  |  | 0.03* |
| Present | 471 (17.1) | 232 (16.8) | 189 (18.2) | 50 (14.9) | 10.8<br>(4) |  |
| Absent | 2152 (78.1) | 1068 (77.4) | 806 (77.6) | 278 (82.7) |  |  |
| Unknown | 131 (4.8) | 80 (5.8) | 43 (4.1) | 8 (2.4) |  |  |
| APOE Genotype |  |  |  |  | - | 0.08 |
| E3/E3 | 992 (36.0) | 500 (36.2) | 345 (33.2) | 147 (43.8) |  |  |
| E3/E4 | 952 (34.6) | 461 (33.4) | 380 (36.6) | 111 (33.0) |  |  |
| E3/E2 | 154 ( 5.6) | 79 ( 5.7) | 57 (5.5) | 18 (5.4) |  |  |
| E4/E4 | 270 ( 9.8) | 143 (10.4) | 106 (10.2) | 21 (6.2) |  |  |
| E4/E2 | 67 (2.4) | 32 (2.3) | 25 (2.4) | 10 (3.0) |  |  |
| E2/E2 | 3 (0.1) | 2 (0.1) | 1 (0.1) | 0 (0.0) |  |  |
| Missing/Unknown | 316 (11.5) | 163 (11.8) | 124 (11.9) | 29 (8.6) |  |  |
| Co-Occurrence of Vascular Dementia |  |  |  |  | 4.2<br>(2) | 0.13 |
| Present | 320 (11.6) | 162 (11.7) | 109 (10.5) | 49 (14.6) |  |  |
| Absent | 2434 (88.40) | 1213 (88.3) | 929 (89.5) | 287 (85.4) |  |  |
| Co-Occurrence of Lewy Body Dementia |  |  |  |  | 0.44<br>(2) | 0.80 |
| Present | 90 (3.3) | 44 (3.2) | 33 (3.2) | 13 (3.9) |  |  |
| Absent | 2664 (96.70) | 1336 (96.8) | 1005 (96.8) | 323 (96.1) |  |  |
| Co-Occurrence of Frontotemporal Dementia |  |  |  |  | - | 0.45 |
| Present | 23 (0.80) | 9 (0.7) | 10 (1.0) | 4 (1.2) |  |  |
| Absent | 2731 (99.20) | 1371 (99.3) | 1028 (99.0) | 332 (98.8) |  |  |

Continuous factors reported as mean (SD) and categorical factors reported as n (%). \* denotes a p value of <0.05. \*\* Differences between clusters evaluated using Chi-square or Fisher's exact test for categorical variables and one-way ANOVA for quantitative variables. Chi-square ( $\chi^2$ ) or correlation ratio (Eta2) reported when applicable.

To determine the significance of problem history on the subsequent cognitive decline of the NACC participants we modeled the longitudinal Clinical Dementia Rating Sum of Boxes (CDRSUM) scale over the next two annual follow-up visits (Figure 2). Problem history subtypes had significantly different changes in CDRSUM while controlling for age, sex, *APOE* genotype, baseline CDRSUM, and visit number (p = 0.013). At visit three, the substance use subtype had the lowest adjusted mean (±SE) change in CDRSUM compared to baseline of 1.51±0.07. The cardiovascular problem history subtype had the highest adjusted mean (±SE) change in CDRSUM of 1.96±0.l6. Post-hoc analyses indicated that these two subtypes were significantly different at visit 3 (p = 0.005) (Figure 2; Supplementary Table 1).

**Figure 2:**
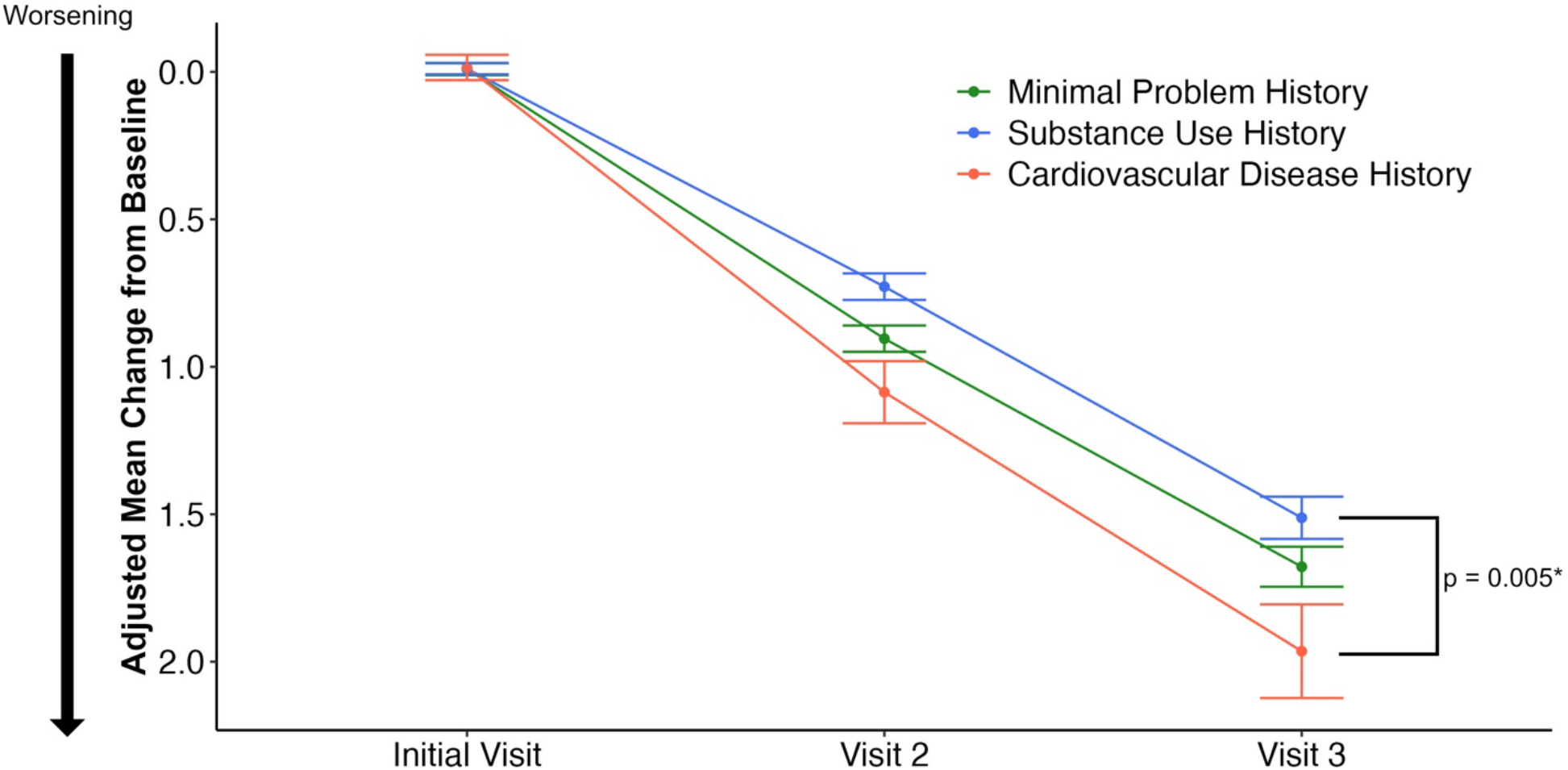
Change in CDR Sum of Boxes for each Problem History Cluster. Change in the Clinical Dementia Rating Sum of Boxes (CDRSUM) in each problem history cluster. Total scores range from 0-18 with higher numbers indicating worsening cognitive decline. Data are plotted as the adjusted mean CDRSUM (± standard error) change from baseline. Data are adjusted using a linear mixed effects model with age, sex, *APOE* genotype, and baseline CDRSUM included as fixed effects and subject ID included as a random effect. *p value corresponds to pairwise comparisons between the marginal mean of each cluster at visit 3.

## 4. Discussion

In this study, we leveraged the NACC database to perform clustering of medical and substance use history (i.e., problem history) in a cohort of participants who developed incident Alzheimer’s Disease at a follow-up visit. We identified three unique problem history subtypes: minimal problem history, substance use history, and cardiovascular problem history. Notably, cognitive decline among problem history subtypes varied with the level of effect being clinically significant over a subsequent follow-up of two years. The difference between the adjusted mean change in CDRSUM between the substance use history and cardiovascular problem history subtype at visit three was 0.45. This is the same difference that was seen between the treatment and placebo groups at 18 months of follow-up in the phase III lecanemab trial [9]. Thus, heterogeneity in problem history among clinic trial participants is likely a significant factor in clinical trials. Future clinical trials should therefore consider problem history in their inclusion/exclusion criteria and their analysis.

Furthermore, our AD clustering study expands on findings from other studies that have clustered on EHR-derived problem history variables in patients already diagnosed with AD. Notably, all three previous clustering studies found a group with higher prevalence of cardiovascular disease—a finding corroborated by our study [28,30,31]. Moreover, we showed a significantly faster decline in cognition over a two-year follow-up period in the cardiovascular problem history subtype. Cardiorespiratory fitness has been observed as a mediating factor in AD treatment [45] and AD progression [46]. Future clinical trials should therefore either specifically target these individuals or account for a cardiovascular disease history to minimize the chance of a type II error. Alexander et al., found a subtype of AD patients with higher rates of smoking that was also associated with more depression and anxiety diagnoses and a faster rate of AD progression [31]. Our study, however, detected a substance use history cluster with no significant difference in rates of depression and a slower cognitive decline over two years. Although depression and substance use disorders are often comorbid [47,48], other reports have found conflicting results in different sexes [49] and ages [50] which could account for our different results. It is also possible that our study was not sensitive enough to characterize depression and anxiety histories given that the data we analyzed included only two questions pertaining to mental health (Table 2). Finally, our study identified a novel minimal problem history cluster with a higher proportion of female and Hispanic individuals. This group of individuals is historically understudied in healthcare [51] and the prevalence of AD in this population is expected to grow by an estimated 460% by 2060 [1]—highlighting the need for future studies to target these individuals.

Our study has a few limitations. First, the NACC cohort analyzed in this study was comprised primarily of highly educated white individuals and thus may not as generalizable as studies using the EHR. However, one advantage of our NACC cohort is that all participants are seen at specialist memory centers and thus likely have more accurate diagnoses. By contrast, EHR-derived data are limited by the issue that AD is often misdiagnosed outside of specialist memory centers [52,53] and that co-pathology can make it difficult to distinguish between different types of dementia [54]. Second, our inclusion criteria targeted individuals who did not have a diagnosis of AD but would receive one in the future—thus the estimated cognitive decline over a two-year period is not generalizable to a wide clinic population of older adults with early cognitive decline. However, our cohort is similar to individuals who would be targeted for a late-phase clinical trial and thus our results remain very relevant to future AD intervention strategies [55]. Third, we were limited to the 26 problem history variables in the NACC questionnaire. Future studies should include additional problem history variables such as generalized anxiety disorder, sleep disorders, kidney disease, autoimmune conditions, etc.

In conclusion, we found three problem history subtypes in a clinical cohort of AD patients: minimal problem history, substance use history, and cardiovascular problem history. The minimal problem history cluster had a higher proportion of Hispanic females and earlier onset AD. The substance use history subtype was comprised primarily of individuals who had smoked more than one hundred cigarettes over their lifetime and had the slowest decline in cognition over a two-year follow-up period. The cardiovascular problem history subtype was older than the other clusters and had a higher proportion of white male individuals. This subtype also had a significantly higher decline in cognition over a two-year follow-up period—indicating results from our study may be informative to future work aimed at designing precision clinical trials.

## Data Availability

This study used data publicly available at the National Alzheimer’s Coordinating Centers (naccdata.org)

https://naccdata.org

## Acknowledgements/Conflicts/Funding Sources/Consent Statement

## Acknowledgements

RAH and OJV conceived the presented idea with input from MES and RT. CM and OJV designed the analysis plan with critical feedback from MES. CM and OJV performed data quality control procedures, conducted clustering analysis, and drafted the initial manuscript. RAH developed the approach for defining maternal family history. RS, RT, RAH, and DRM provided expertise regarding the analysis and interpretation of findings. All authors revised, discussed, and approved the final manuscript.

## Consent Statement

Informed consent was not necessary for this study because all data were de-identified.

