## Supplementary Figure 1 for "Distinct medical and substance use histories associate with cognitive decline in Alzheimer’s Disease"

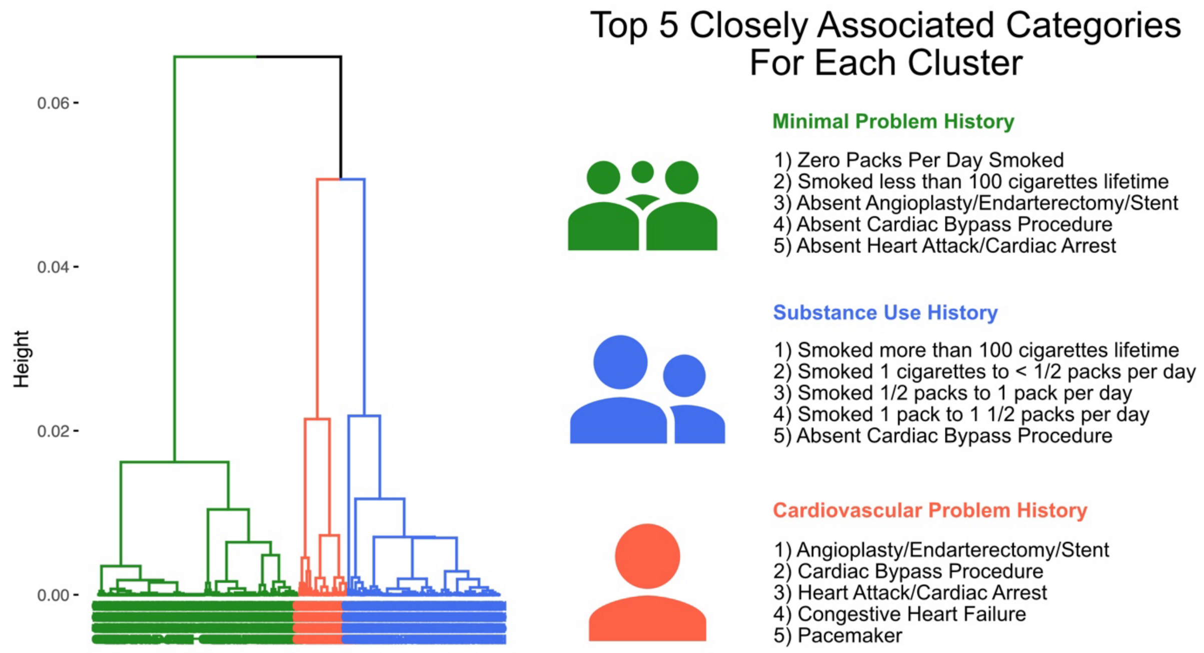


**Supplementary Figure 1: Dendrogram from Clustering Analysis.** The height of each branch represents the relationship between individuals in the dataset. The top five most closely associated problem history items are shown and were used to inform the naming of each cluster.
