## Supplementary Table 1 for "Distinct medical and substance use histories associate with cognitive decline in Alzheimer’s Disease"

**Supplementary Table 1. Pairwise Comparisons of Problem History Clusters on Clinical Dementia Rating Sum of Boxes at NACC Visit 3**

| | Contrast (95% CI) | p value |
|---|---|---|
| Minimal – Substance Use | 0.11 (-0.01 - 0.23) | 0.067 |
| Minimal – Cardiovascular History | -0.15 (-0.32 - 0.03) | 0.098 |
| Substance Use – Cardiovascular History | -0.25 (-0.43 - -0.08) | 0.005 |

Pairwise comparisons between each problem history cluster on the Clinical Dementia Rating Sum of Boxes (CDRSUM) at visit 3 adjusting for age, sex, *APOE* genotype, baseline CDRSUM, and visit number. The reported estimates reflect the adjusted mean difference in CDRSUM between clusters with 95% confidence intervals and p values representing statistical significance.
